# Serum anion gap is associated with all-cause mortality among critically ill patients with congestive heart failure

**DOI:** 10.1101/2020.07.24.20161463

**Authors:** Yiyang Tang, Wenchao Lin, Lihuang Zha, Xiaofang Zeng, Zhenghui Liu, Zaixin Yu

## Abstract

**Background:** Congestive heart failure (CHF) is a complex clinical syndrome, with high morbidity and mortality. Serum anion gap (SAG) has been known to be associated with the severity of various cardiovascular diseases. However, the role of SAG indicators in CHF is unclear.

**Methods and results:** A retrospective analysis of data from MIMIC-III v1.4 was conducted in critically ill patients with CHF. Clinical information of each patient, including demographic data, comorbidities, vital signs, scores, and laboratory indicators, were successfully obtained. Cox proportional hazards models were performed to determine the relationship between SAG and mortality in CHF patient, the consistency of which was further verified by subgroup analysis.

**Results:** A total of 7426 subjects met the inclusion criteria. In multivariate analysis, after adjusting for age, gender, ethnicity, and other potential confounders, higher SAG was significantly related to an increase in 30-day and 90-day all-cause mortality of critically ill patients with CHF compared with lower SAG (tertile3 vs tertile1: adjusted HR, 95% CI: 1.74, 1.46–2.08; 1.53, 1.32–1.77). In subgroup analysis, the association between SAG and all-cause mortality present similarities in most strata.

**Conclusion:** SAG at admission can be a promising predictor of all-cause mortality in critically ill patients with CHF.

## 1 Introduction

Congestive heart failure (CHF) is the end-stage manifestation of various cardiovascular diseases with structural and functional disruptions in the myocardium, which causes restricted ventricular ejection or filling, systemic blood circulation disorder, insufficient perfusion of tissues and organs, and ultimate death ^[1]^. CHF has become a major public health problem with more than 23 million patients affected worldwide and it still has a high mortality despite great progress in diagnosis and treatment for the last few decades ^[2]^. Thus, how to identify high-risk patients among CHF early, determine their prognosis, and formulate effective individualized interventions has attracted more and more attention from clinicians in recent years.

Metabolic acidosis is a common complication of CHF, which is closely related to ischemia and hypoxia of tissue caused by hemodynamic disorders and the use of diuretics ^[3]^. In turn, acidosis and accompanying hyperkalemia can further weaken myocardial contractility, creating a vicious circle. Acidosis serves as the independent predictor for the long-term prognosis of CHF and pH value could help to stratified the risks ^[4]^. Nevertheless, the predictive value of other laboratory parameters that reflect acid-base imbalance in CHF requires more evidence, such as serum anion gap (SAG).

SAG refers to the difference between undetermined anions and cations, indicating the concentration of fixed acids in plasma, and is a commonly used and easily obtained laboratory parameter of acid-base imbalance ^[5]^. Some recent studies have confirmed that SAG is elevated and closely associated with the poor prognosis of a variety of diseases, including acute pesticide poisoning ^[6]^, sepsis ^[7]^, acute and chronic kidney injury ^[8, 9]^, trauma ^[10]^, and coronary artery disease ^[11, 12]^. However, it still remains unclear whether the anion gap can be used as a prognostic marker for the critically ill patients with CHF. Therefore, we conducted this study with the aim of investigating the association between SAG and the mortality in these patients.

## 2 Materials and Methods

### 2.1 Data source

The Multiparameter Intelligent Monitoring in Intensive Care III version 1.4 (MIMIC-III v1.4) is a freely accessible critical care database developed and operated by the Massachusetts Institute of Technology, which contains the detailed clinical data of 53423 adult patients (age more than 16 years) from June 2001 to October 2012 in the intensive care units of Beth Israel Deaconess Medical Center ^[13]^. Before implementing this research, author Tang completed and passed the CITI “Data or Specimens Only Research” course (No.9014457) and obtained authorization for database access. In need of special note here is that this database was approved by the institutional review boards of Massachusetts Institute of Technology (Cambridge, MA, USA) and Beth Israel Deaconess Medical Center (Boston, MA, USA), and no additional ethical approval need to be provided here.

### 2.2 Study Population and Design

In the MIMIC-III database, all adult patients aged over 18 years old, first admitted to the intensive care unit (ICU) and diagnosed with CHF were included in the study, according to according to ninth revision of the international classification of diseases code (code = 4280). Patients with any of the following criteria were excluded from the study: (1) ICU stay time is less than 24 hours (2) no anion gap results within 24 hours of admission to the ICU (3) survival time is less than 0 (some organ donors may die earlier than admission). The workflow was shown in Figure 1.

**Figure 1.**
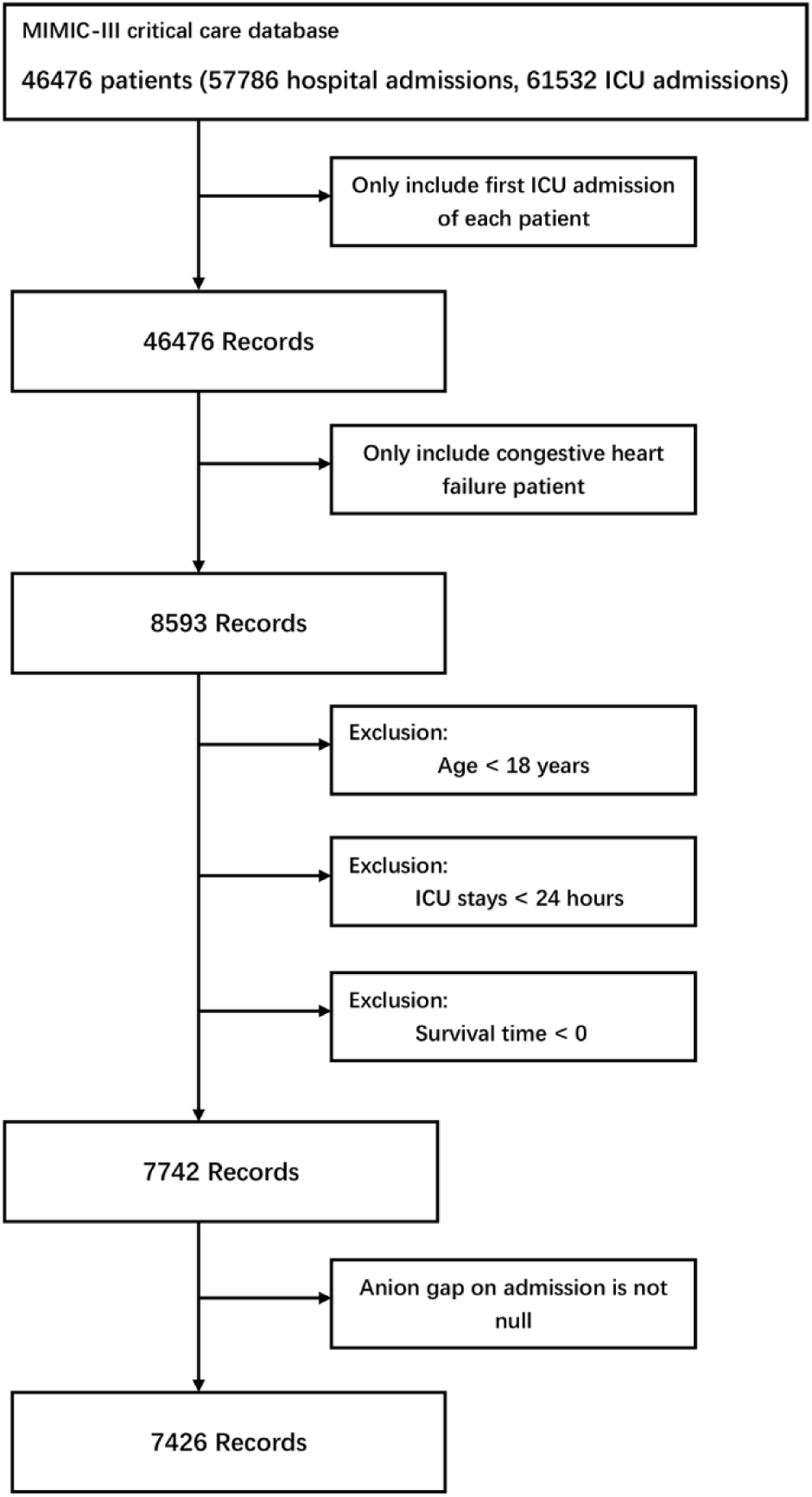
Workflow of the data extraction. The inclusion and exclusion criteria of the study subjects were revealed, and 7426 subjects were finally included. ICU, intensive care unit.

The starting point of the study was defined as the time of admission to the ICU for patients with CHF, and the end point was the time of death, 30 or 90 days after admission. The patients’ 30-day and 90-day mortality was chosen as the primary outcome of interest and death data was extracted from the Social Security Death Index.

### 2.3 Data Extraction and Preparation

We utilized structured query language (SQL) to extract clinical data with PostgreSQL tools (version 9.6), including demographics, comorbidities, vital signs, severity scores, laboratory tests, interventions, and others. The demographics included age, gender, and ethnicity. To protect the privacy of patients, the database has had the date of birth shifted to exactly 300 years for patients who are older than 89 years old. Before the analysis, the age of these patients was adjusted according to the following formula: real age=age-300+89 ^[14]^. The vital sign contains temperature, respiratory rate (RR), heart rate (HR), systolic blood pressure (SBP), diastolic blood pressure (DBP), mean blood pressure (MBP), percutaneous oxygen saturation (SpO_2_). The comorbidities included acute myocardial infarction (AMI), atrial fibrillation, valvular heart diseases, pulmonary circulation diseases, hypertension (HBP), diabetes, pneumonia, respiratory failure, liver disease, renal failure, stroke, and malignancy. Calculate the sequential organ failure assessment score (SOFA) and simplified acute physiology score II (SAPSII) when entering the ICU for each patient to assess the severity of the disease ^[15, 16]^. The laboratory tests within 24 hours after ICU admission were also extracted, including SAG, white blood cell (WBC), platelet, hemoglobin, blood urea nitrogen (BUN), creatinine, sodium, potassium, chloride, bicarbonate, glucose, prothrombin time (PT), activated partial thromboplastin time (APTT), lactate, plasma N-terminal pro brain natriuretic peptide (NT-proBNP). The interventions included vasopressor, dialysis, and mechanical ventilation.

After extracted, the raw data was merged and disposed with patient identifiers by using STATA version 16 (https://www.stata.com/). The “winsorize” function was used to reduce the effect of outliers and multivariate multiple imputation with chained equations was utilized to impute the missing values. The details of missing value were shown in Table 1, and we noticed that more than 20% of the subjects did not have a record of lactate and NT-proBNP in our cohort, which was converted and considered as a dummy variable in our models to avoid the possible bias caused by direct filling of missing value ^[17]^.

**Table 1.** Details of missing values.

| Variables | The number of missing values | The percent of missing values | Variables | The number of missing values | The percent of missing values |
| --- | --- | --- | --- | --- | --- |
| SBP | 39 | 0.5% | glucose | 48 | 0.6% |
| DBP | 39 | 0.5% | lactate | 1993 | 26.8% |
| MBP | 32 | 0.4% | NT-proBNP | 6231 | 83.9% |
| RR | 37 | 0.5% | PT | 111 | 1.5% |
| HR | 32 | 0.4% | APTT | 137 | 1.8% |
| SpO<sub>2</sub> | 34 | 0.5% | sodium | 1 | <0.1% |
| temperature | 255 | 3.4% | potassium | 1 | <0.1% |
| weight | 484 | 6.5% | chloride | 1 | <0.1% |
| WBC | 68 | 0.9% | bicarbonate | 2 | <0.1% |
| platelet | 65 | 0.8% | creatinine | 2 | <0.1% |
| hemoglobin | 88 | 1.1% | BUN | 4 | <0.1% |

Note: SBP, systolic blood pressure; DBP, diastolic blood pressure; MBP, mean blood pressure; RR, respiratory rate; HR, heart rate; SpO<sub>2</sub>, percutaneous oxygen saturation; WBC, white blood cell; BUN, blood urea nitrogen; PT, prothrombin time; APTT, activated partial thromboplastin time; NT-proBNP, N-terminal pro brain natriuretic peptide.

### 2.4 Statistical Analysis

The baseline data of all subjects are divided into three groups by SAG tertiles. The continuous variables were presented as mean ± SD or medians and interquartile range (IQR). Kruskal Wallis test was used to conduct hypothesis test for continuous variables. The categorical variables are expressed by numbers and percentages, which were analyzed by Chi-square (or Fisher’s exact) tests. A generalized additive model (GAM) was used to determine the nonlinear association between SAG and 30-day and 90-day all-cause mortality in the critically ill patients with CHF. In addition, we visually shown the relationship between SAG and the survival in CHF patients through the Kaplan-Meier (K-M) curve, and tested it through Log-Rank test.

To determine the independent prognostic value of SAG for critically ill patients with CHF, we performed the Cox proportional hazard model to analyze the relationship between SAG and 30-day and 90-day all-cause mortality in these patients, using the first tertile or quartile as the reference and adjusting for potential confounders. Two multivariate models were conducted and the result were described as hazard ratios (HRs) with 95% confidence intervals (CIs). In model I, covariates were only adjusted for the confounders age, sex, and ethnicity. On the basis of model I, model II further adjusted for other confounders including temperature, HR, RR, SBP, DBP, SpO_2_, weight, atrial fibrillation, liver disease, valvular heart diseases, pulmonary circulation diseases, pneumonia, respiratory failure, diabetes, stroke, malignancy, SOFA, SAPSII, lactate, NT-proBNP, PT, WBC, BUN, creatinine, potassium, bicarbonate, glucose, vasopressor, dialysis, and mechanical ventilation. We choose these factors as confounders based on their association with the outcomes or a change in effect estimate exceeding 10% ^[18]^. Besides, a subgroup analysis of the correlation between SAG and 30-day all-cause mortality was performed to examine whether the impact of SAG in various subgroups was different. All statistical analyses were performed on EmpowerStats version 2.20 (http://www.empowerstats.com/cn/, X&Y solutions, Inc., Boston, MA) and R software version 3.4.3; P < 0.05 (2-sided) was considered statistically significant.

## 3 Results

### 3.1 Clinical Characteristics of Subjects

In this retrospective study, there were a total of 7426 subjects, 3971 of whom were male and 3455 were female. The age of the subjects was generally older, with a median of 75.3 years old. The subjects were mostly white, accounting for 73.3 percent share of the total. And the 30-day and 90-day overall mortality was 17.7% and 24.4%, respectively. The clinical characteristics of these subjects stratified by SAG tertiles were shown in Table 2. A total of 2334 subjects were in the low-SAG group (tertile 1, SAG < 13), 2519 subjects were in the mid-SAG group (tertile 2, SAG≥ 13 and SAG < 16), and 2573 subjects were in the high-SAG group (tertile 3, SAG ≥16). Subjects with higher SAG levels had more comorbidities of HBP, AMI, diabetes, renal failure, and respiratory failure, with higher mortality, higher SOFA and SAPSII scores, and higher rates of use of vasopressor, dialysis, and mechanical ventilation.

**Table 2.** The clinical characteristics of critically ill patients with CHF according to SAG levels.

| Characteristics | Serum anion gap (mmol/L) |  |  | P value |
| --- | --- | --- | --- | --- |
|  | <13 (n=2334) | ≥13, <16 (n=2519) | ≥16 (n=2573) |  |
| Age (years) | 72.2±13.2 | 73.2±13.2 | 72.5±13.5 | 0.013 |
| Gender, n (%) |  |  |  | 0.278 |
| Male | 1280 (54.8) | 1332 (52.9) | 1359 (52.8) |  |
| Female | 1054 (45.2) | 1187 (47.1) | 1214 (47.2) |  |
| Ethnicity, n (%) |  |  |  | 0.002 |
| White | 1744 (74.7) | 1881 (74.7) | 1821 (70.8) |  |
| Black | 152 (6.5) | 187 (7.4) | 230 (8.9) |  |
| Other | 438 (18.8) | 451 (17.9) | 522 (20.3) |  |
| SBP, mmHg | 116.7±15.2 | 117.6±17.0 | 116.2±17.8 | 0.004 |
| DBP, mmHg | 57.3±9.4 | 58.0±10.4 | 58.1±10.9 | 0.013 |
| MBP, mmHg | 75.3±9.7 | 76.1±10.5 | 75.9±11.1 | 0.038 |
| HR, beats/minute | 84.0±14.2 | 84.0±15.6 | 86.8±16.7 | <0.001 |
| RR, beats/minute | 18.7±3.7 | 19.6±3.9 | 20.2±4.3 | <0.001 |
| Temperature, °C | 36.8±0.6 | 36.8±0.6 | 36.7±0.7 | <0.001 |
| SpO<sub>2</sub>, % | 97.3±1.9 | 96.9±2.0 | 96.8±2.4 | <0.001 |
| Weight, Kg | 81.9±24.1 | 81.2±24.3 | 81.3±24.8 | 0.212 |
| Scoring systems |  |  |  |  |
| SOFA | 4.5±2.7 | 4.4±2.8 | 5.7±3.3 | <0.001 |
| SAPSII | 37.1±11.6 | 38.6±12.6 | 44.2±14.0 | <0.001 |
| Dialysis, n (%) | 60 (2.6) | 115 (4.6) | 440 (17.1) | <0.001 |
| Vasopressor, n (%) | 558 (23.9) | 498 (19.8) | 666 (25.9) | <0.001 |
| Ventilation, n (%) | 624 (26.7) | 784 (31.1) | 1000 (38.9) | <0.001 |
| ICU LOS, hours | 116.7±149.8 | 130.3±164.5 | 147.4±180.9 | <0.001 |
| 30-day mortality, n (%) | 254 (10.9) | 399 (15.8) | 665 (25.8) | <0.001 |
| 90-day mortality, n (%) | 411 (17.6) | 579 (23.0) | 882 (34.3) | <0.001 |
|  | <13 (n=2334) | ≥13, <16 (n=2519) | ≥16 (n=2573) |  |
| Comorbidities, n (%) |  |  |  |  |
| Liver diseases | 97 (4.2) | 89 (3.5) | 119 (4.6) | 0.144 |
| Renal failure | 325 (13.9) | 552 (21.9) | 813 (31.6) | <0.001 |
| Atrial fibrillation | 1053 (45.1) | 1134 (45.0) | 1116 (43.4) | 0.377 |
| Stroke | 105 (4.5) | 152 (6.0) | 134 (5.2) | 0.056 |
| AMI | 121 (5.2) | 207 (8.2) | 270 (10.5) | <0.001 |
| VHD | 178 (7.6) | 248 (9.8) | 264 (10.3) | 0.003 |
| PCD | 130 (5.6) | 140 (5.6) | 139 (5.4) | 0.959 |
| HBP | 278 (11.9) | 467 (18.5) | 673 (26.2) | <0.001 |
| Diabetes | 786 (33.7) | 846 (33.6) | 1021 (39.7) | <0.001 |
| Pneumonia | 440 (18.9) | 581 (23.1) | 640 (24.9) | <0.001 |
| Respiratory failure | 487 (20.9) | 628 (24.9) | 774 (30.1) | <0.001 |
| Malignancy | 94 (4.0) | 145 (5.8) | 118 (4.6) | 0.015 |
| Laboratory tests |  |  |  |  |
| WBC (K/ul) | 11.4±6.2 | 12.7±27.8 | 13.8±8.3 | <0.001 |
| Platelet (K/ul) | 196.2±95.4 | 218.7±98.2 | 240.1±118.0 | <0.001 |
| Hemoglobin (g/dl) | 10.2±1.9 | 10.8±2.0 | 10.9±2.1 | <0.001 |
| Creatinine (mg/dl) | 1.0±0.6 | 1.4±0.9 | 2.4±2.0 | <0.001 |
| BUN (mg/dl) | 23.2±13.7 | 30.0±19.0 | 45.1±28.8 | <0.001 |
| Glucose (mg/dl) | 129.0±53.7 | 144.7±60.9 | 171.3±94.8 | <0.001 |
| Sodium (mmol/L) | 138.8±4.6 | 138.9±4.6 | 138.2±5.4 | <0.001 |
| Potassium (mmol/L) | 4.1±0.6 | 4.2±0.7 | 4.4±0.8 | <0.001 |
| Chloride (mmol/L) | 106.2±6.2 | 104.6±5.9 | 102.7±6.6 | <0.001 |
| Bicarbonate (mmol/L) | 26.5±4.9 | 24.8±4.2 | 21.5±4.7 | <0.001 |
| PT (second) | 15.9±7.1 | 16.7±9.9 | 17.8±12.0 | <0.001 |
|  | <13 (n=2334) | ≥13, <16 (n=2519) | ≥16 (n=2573) |  |
| Laboratory tests |  |  |  |  |
| APTT (second) | 38.7±23.8 | 38.8±25.2 | 40.4±26.6 | 0.002 |
| Lactate |  |  |  | <0.001 |
| <1.7 mmol/L | 1076 (46.1) | 865 (34.3) | 694 (27.0) |  |
| ≥1.7 mmol/L | 634 (27.2) | 858 (34.1) | 1306 (50.8) |  |
| No test | 624 (26.7) | 796 (31.6) | 573 (22.3) |  |
| NT-proBNP |  |  |  | <0.001 |
| <4983 pg/ml | 192 (8.2) | 219 (8.7) | 186 (7.2) |  |
| ≥4983 pg/ml | 118 (5.1) | 200 (7.9) | 280 (10.9) |  |
| No test | 2024 (86.7) | 2100 (83.4) | 2107 (81.9) |  |

Note: CHF, congestive heart failure; SAG, serum anion gap; SBP, systolic blood pressure; DBP, diastolic blood pressure; MBP, mean blood pressure; RR, respiratory rate; HR, heart rate; SpO<sub>2</sub>, percutaneous oxygen saturation; SOFA, stroke, and malignancy. Calculate the sequential organ failure assessment score; SAPSII, simplified acute physiology score II; ICU, intensive care unit; LOS, length of stay; AMI, acute myocardial infarction; VHD, valvular heart diseases; PCD, pulmonary circulation diseases; HBP, hypertension; WBC, white blood cell; BUN, blood urea nitrogen; PT, prothrombin time; APTT, activated partial thromboplastin time; NT-proBNP, N-terminal pro brain natriuretic peptide.

### 3.2 Association between SAG and all-cause mortality

As shown in Figure 2, the result of GAM analysis indicated that there was a U-shaped relationship between SAG and 30-day all-cause mortality in critically ill patients with CHF. K-M survival curve illustrated that subjects with higher SAG levels presented lower survival rate and shorter survival time (Log-Rank p < 0.0001, Figure 3).

**Figure 2.**
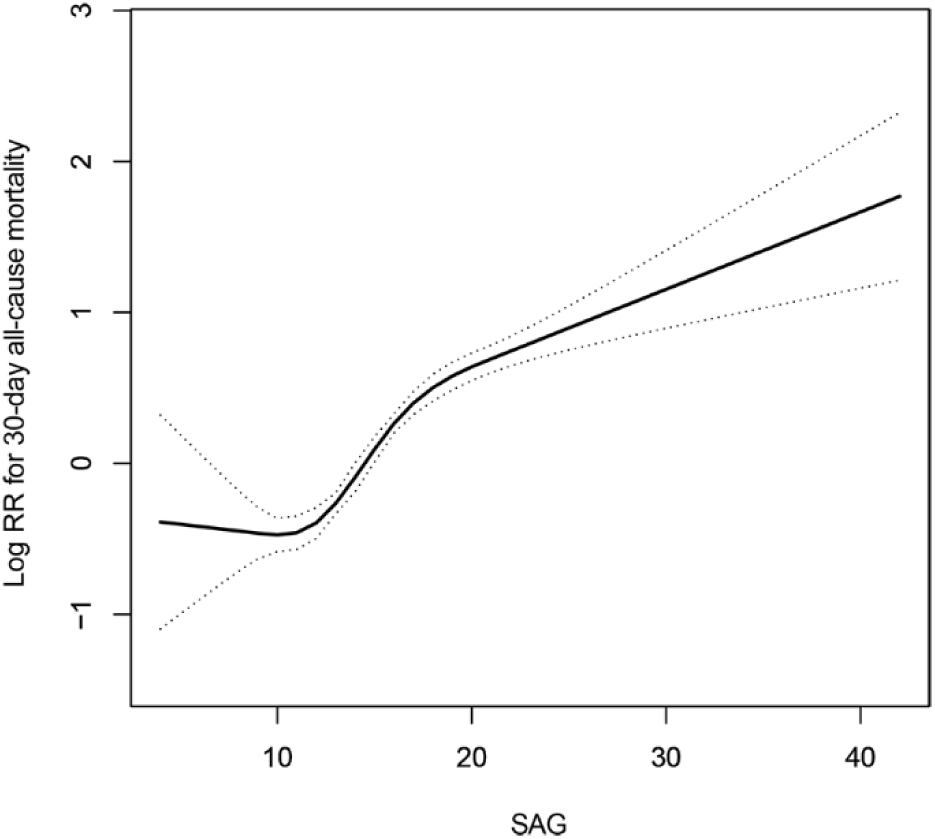
Construction of the smooth curve fitting of the risk of 30-day all-cause mortality and SAG using a generalized additive model. Dashed curves were for the 95% of confidence interval.

**Figure 3.**
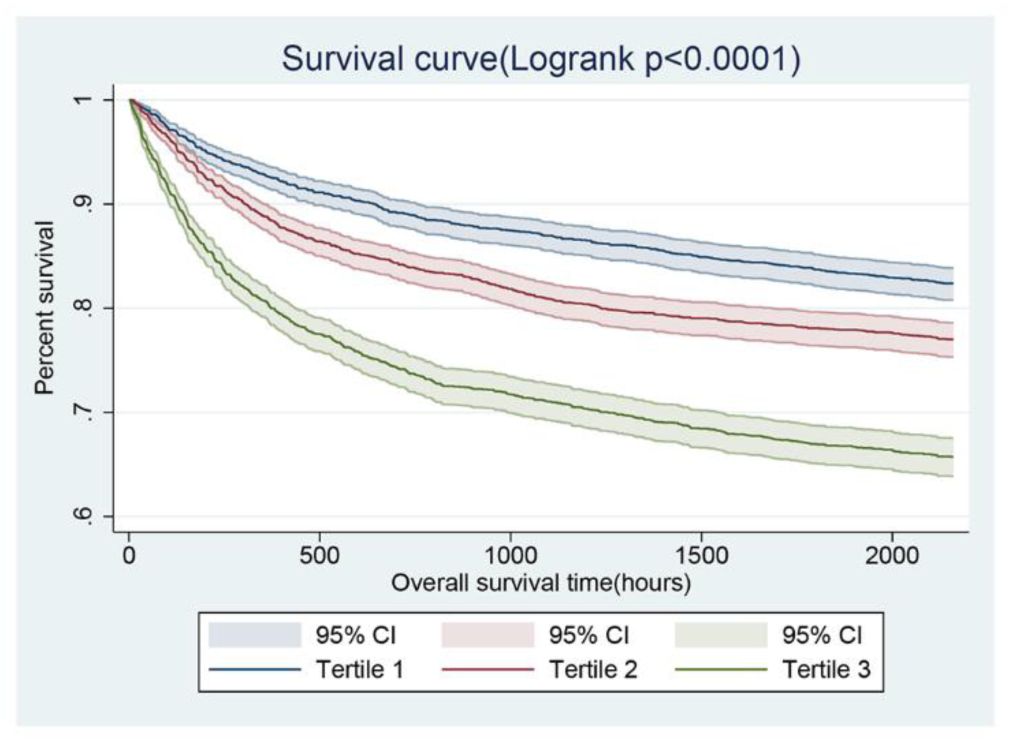
Kaplan-Meier survival curves for critically ill patients with CHF based on tertile of SAG. X-axis: survival time (hours). Y-axis: cumulative survival probability. CHF, congestive heart failure. SAG, serum anion gap.

The Cox proportional hazard model was used to assess the association between SAG and all-cause mortality and presented in Table 3. In model I, after adjusting for the confounders age, sex, and ethnicity, high levels of SAG were significantly associated with increased risk of 30-day and 90-day all-cause mortality (tertile 3 versus tertile 1: HR, 95% CI: 2.62, 2.27–3.03; 2.23, 1.98–2.51; quartiles versus quartiles 1: HR, 95% CI: 3.20, 2.68–3.82; 2.62, 2.27–3.02). In model II, after adjusting for age, sex, ethnicity, temperature, HR, RR, SBP, DBP, SpO_2_, weight, atrial fibrillation, liver disease, valvular heart diseases, pulmonary circulation diseases, pneumonia, respiratory failure, diabetes, stroke, malignancy, SOFA, SAPSII, lactate, NT-proBNP, PT, WBC, BUN, creatinine, potassium, bicarbonate, glucose, vasopressor, dialysis, and mechanical ventilation, high levels of SAG were still an independent predictor of 30-day and 90-day all-cause mortality (tertile 3 versus tertile 1: HR, 95% CI: 1.74, 1.46–2.08; 1.53, 1.32–1.77; quartiles 4 versus quartiles 1: HR, 95% CI: 1.97, 1.59–2.45; 1.66, 1.39–1.98).

**Table 3.**
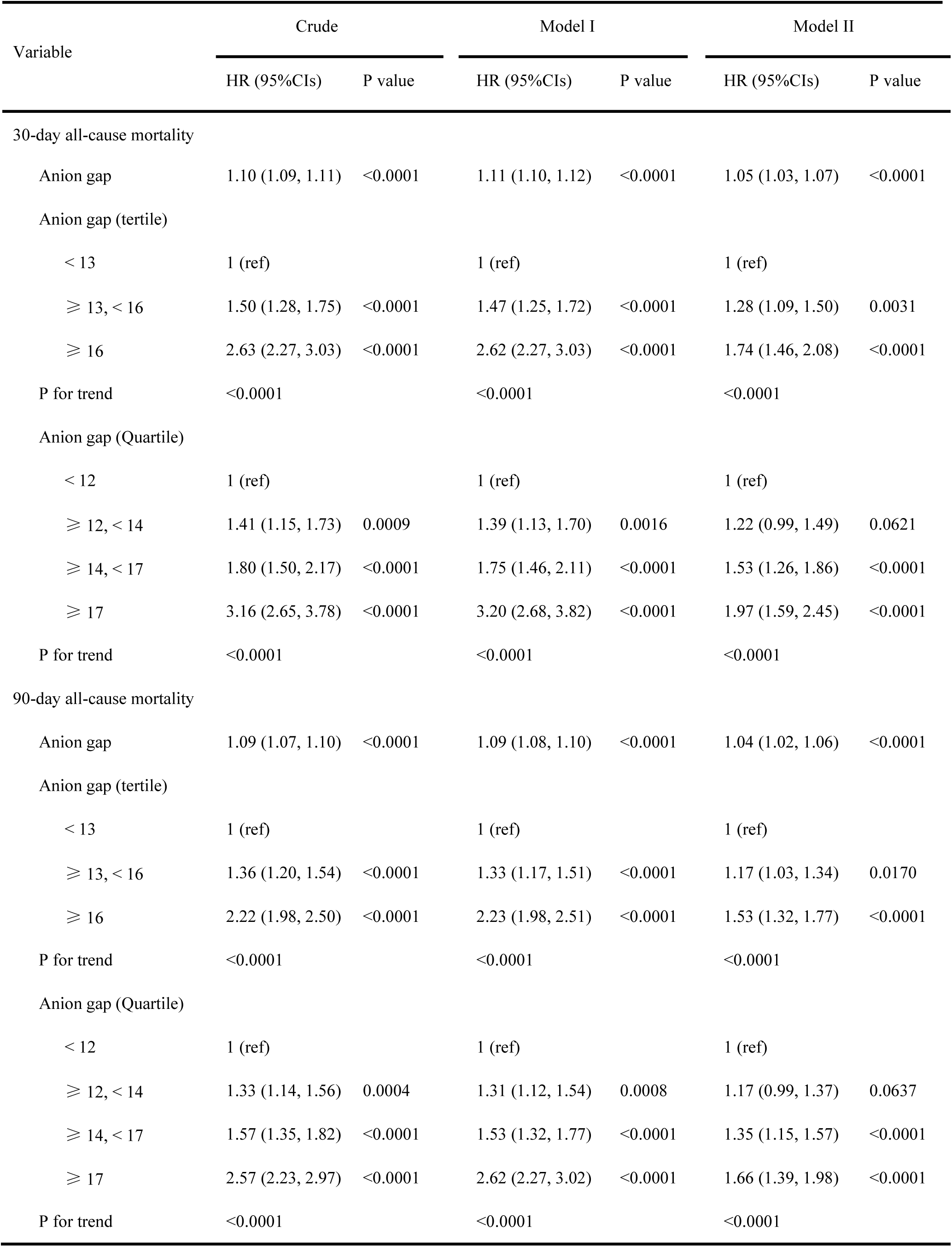
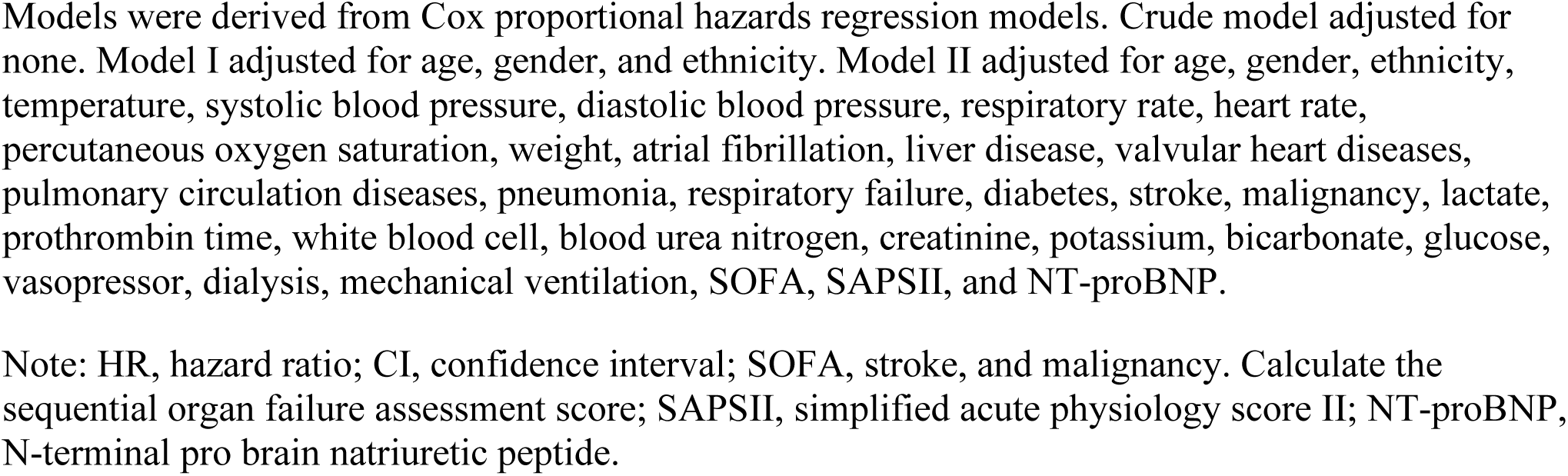
HRs (95% CIs) for all-cause mortality across groups of serum anion gap.

### 3.3 Subgroup analyses

Subgroup analyses were employed to assess the association between the SAG and 30-day all-cause mortality, and there were no significant interactions in most strata as shown in Table 4, except for pneumonia, malignancy, respiratory failure, and mechanical ventilation. Among CHF patients with high SAG, patients with a comorbidity of pneumonia had a significant lower 30-day mortality risk (HR, 95% CI: 1.44, 1.13–1.85 versus 3.27, 2.73–3.91). Similar trends also appeared in patients with a history of malignancy, respiratory failure, and mechanical ventilation (HR, 95% CI: 1.84, 1.10–3.07 versus 2.71, 2.33–3.15; 1.49, 1.22–1.83 versus 3.51, 2.85–4.32; 1.89, 1.55–2.30 versus 2.95, 2.38– 3.65).

**Table 4.**
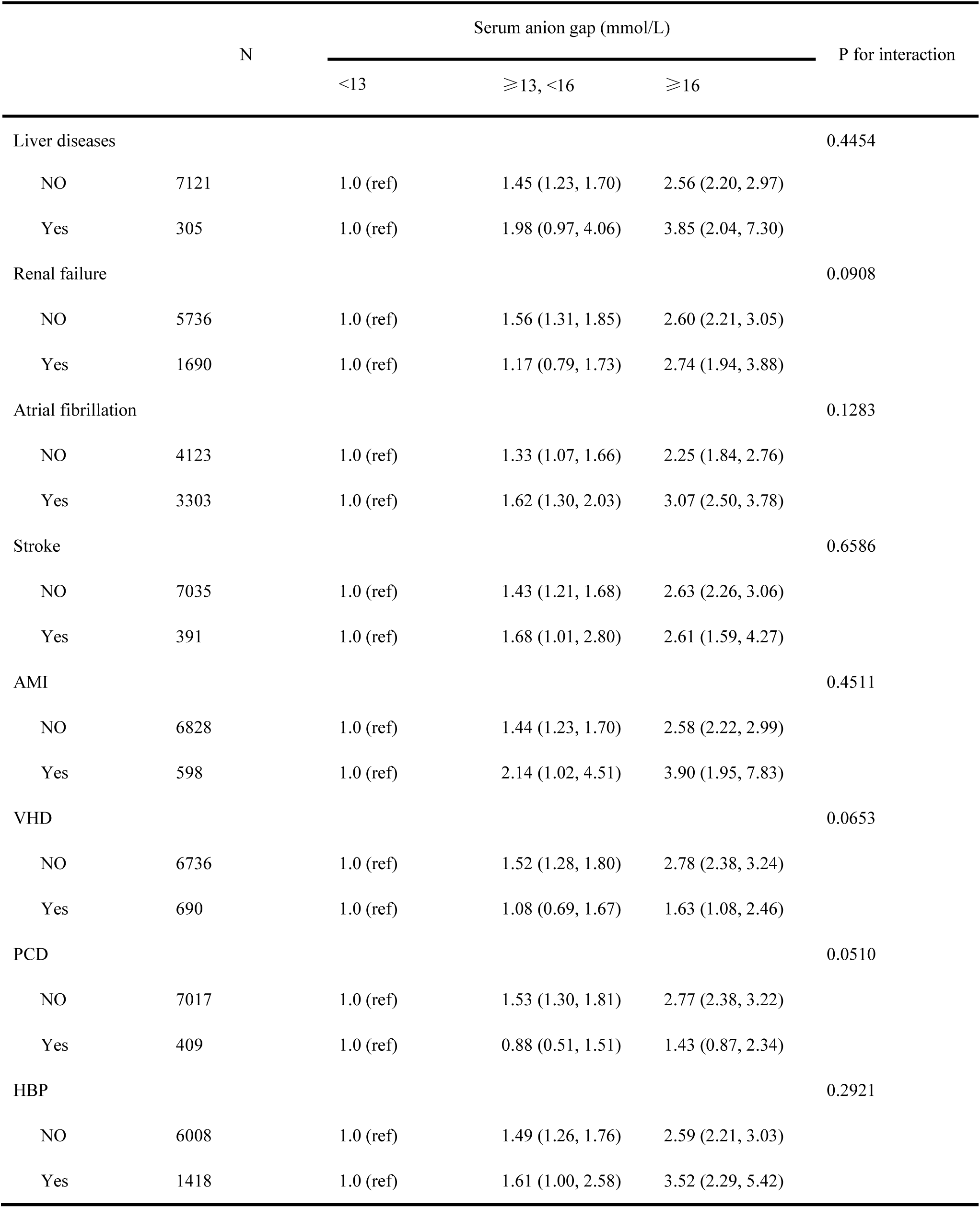
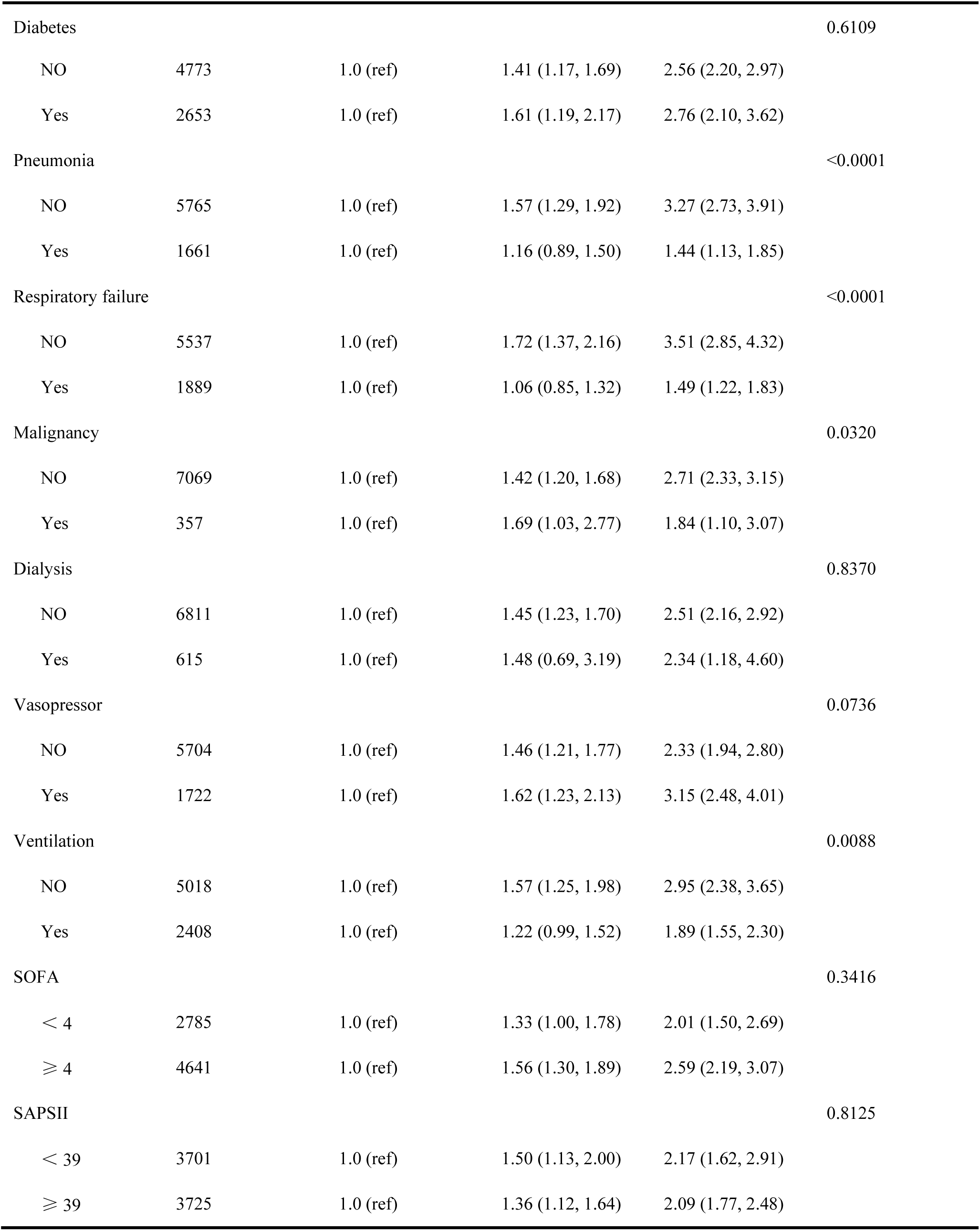
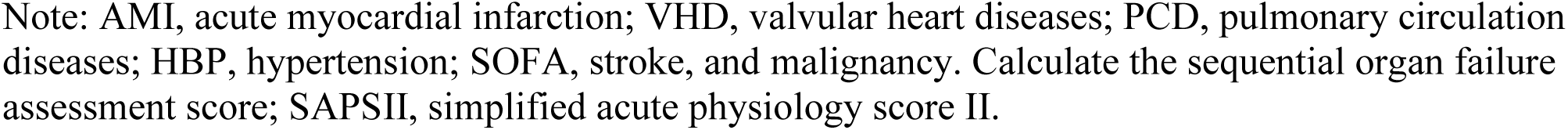
Subgroup analysis of the correlation between serum anion gap and 30-day all-cause mortality in critically ill patients with congestive heart failure.

## 4 Discussion

CHF is a common critical illness in the field of cardiovascular disease, with the characteristics of high prevalence, hospital admissions, including readmissions, and even mortality ^[19]^. According to the Framingham study, the 5-year survival rate of CHF patients for men is only 25%, and that for women is 38%, which heavily threatens human health ^[20, 21]^. Unfortunately, there is a lack of clinical objective indicators to predict the prognosis of patients with CHF at present. Clinically, various laboratory tests such as blood gas analysis and biochemical testing are often used to determine the status of acid-base balance of patients, but the relationship between these indicators and the prognosis of patients with CHF is rarely studied. Therefore, it is particularly important to find a simple and effective laboratory parameter to predict the prognosis of CHF patients, which is also valuable for clinical treatment.

In the present study, we explored the possible prognostic value of SAG in patients with CHF. The relationship between SAG and 30-day all-cause mortality in critically ill patients with CHF presented a U-shaped curve. Higher SAG was associated with increased risk of 30-day and 90-day all-cause mortality and it remained as an independent predictor for all-cause mortality in critically ill patients with CHF after adjusting age, gender, ethnicity, and another confounder. Besides, there were no significant interactions between SAG and most covariables for 30-day mortality, which enhanced the stability and consistency of our results. To the best of our knowledge, our research is the first to clarify that the high levels of SAG was associated with the poor prognosis of critically ill patients with CHF.

As a routine examination that almost all admitted patients would complete, SAG has the characteristics of simple calculation and easy acquisition, even without arterial puncture ^[14]^. SAG, a novel prognostic biomarker, has broad application prospects, but its specific mechanism is still unclear. In simple terms, SAG refers to the remaining unmeasured anions in the serum, including organic acid anions (lactate, keto acid) and inorganic acid anions (PO_4_^2-^, SO_4_^2-^) ^[22]^. Previous studies have illustrated that the accumulation of serum lactate and ketone body accounts for 62% of the cause of the increase of SAG ^[23]^. In CHF, the heart loses its ability to efficiently pump blood, which leads to decreased perfusion of tissues and hypoxia of cells ^[24]^. Under anaerobic conditions, glucose undergoes glycolysis and eventually generates lactate, which may be primarily responsible for increased SAG in the patients with CHF ^[25]^. In addition, sympathetic excitation in CHF also contributes to the overproduction of lactate ^[26]^. Under normal, the production and clearance of lactate are in balance, and its clearance is mainly responsible for the liver and kidney. Patients with CHF are often complicated by liver and kidney dysfunction, which further aggravates hyperlactataemia and the increase of SAG ^[27]^. We also completed the research about the relationship between lactate and 30-day all-cause mortality in critically ill patients with CHF. Consistent with the results of previous studies^[28]^, the increase of lactate was significantly correlated with the poor prognosis (HR, 95% CI: 1.73, 1.53–1.95). After adjusting for covariates including lactate, the anion gap is still associated with poor prognosis, showing the unique value of SAG as an independent predictor of the prognosis of critically ill patients with CHF.

There are still some limitations in our research. First of all, this study was a single-center study, whose subjects were relatively seriously ill. The results of the study may not be applicable to all CHF patients. Second, there are still some unknown or even vital risk factors that have not been considered, although we had used two multivariate model to control the influence of confounder on the outcome variables. Third, only use the SAG concentration of patients when they entered the ICU to assess the relationship between them and all-cause mortality. It may be more valuable for prognosis prediction if SAG can be dynamically monitored. Last, due to the lack of records of albumin, we did not perform albumin correction on the SAG, although some studies have shown that hypoalbuminemia could affect the SAG concentration ^[29]^.

## 5 Conclusion

SAG can effectively predict the 30-day and 90-day all-cause mortality of critically ill patients with CHF, and is expected to become a simple and effective marker for prognostic evaluation in these patients. Clinically, the monitoring of SAG concentration can help to identify high-risk patients early and choose more scientific treatment plan for patients.

## Data Availability

Publicly available datasets were analyzed in this study. This data can be extracted from Monitoring in Intensive Care Database III version 1.4 (MIMIC-III v.1.4) after passing on the required courses and obtaining the authorization.

https://mimic.physionet.org/

## 6 Author Contributions

Author Z. X. Y designed the research. Y.Y.T performed the data analysis and drafted the manuscript. W.C.L, L.H.Z, X.F.Z, and Z.H.L analyzed the data and revised the manuscript.

## 7 Conflicts of Interest

All authors declare that there is no conflict of interest.

## 8 Acknowledgement

Our study was supported by the National Natural Science Foundation of China (81873416) and the National Science and Technology Major Project (2017ZX0930401405).

## Notes

### Competing Interest Statement

The authors have declared no competing interest.

### Clinical Trial

This study is a retrospective study based on the MIMIC-III v1.4, a freely accessible critical care database.This registration is not mandatory for retrospective studies.

### Author Declarations

The MIMIC-III database has received ethical approval from the institutional review boards (IRBs) at Beth Israel Deaconess Medical Center and Massachusetts Institute of Technology. Because the database does not contain protected health information, a waiver of the requirement for informed consent was included in the IRB approval.

## References

[1] Sun D, Zhang F, Ma T, et al. Atorvastatin alleviates left ventricular remodeling in isoproterenol-induced chronic heart failure in rats by regulating the RhoA/Rho kinase signaling pathway [J]. Pharmacol Rep, 2020. DOI:10.1007/s43440-020-00085-3.

[2] Orso F, Fabbri G, and Maggioni AP. Epidemiology of Heart Failure [J]. Handb Exp Pharmacol, 2017, 243: 15–33. DOI:10.1007/164_2016_74.

[3] Peixoto AJ, and Alpern RJ. Treatment of severe metabolic alkalosis in a patient with congestive heart failure [J]. Am J Kidney Dis, 2013, 61(5):822–827. DOI:10.1053/j.ajkd.2012.10.028.

[4] Park JJ, Choi DJ, Yoon CH, et al. The prognostic value of arterial blood gas analysis in high-risk acute heart failure patients: an analysis of the Korean Heart Failure (KorHF) registry [J]. Eur J Heart Fail, 2015, 17(6):601–611. DOI:10.1002/ejhf.276.

[5] Glasmacher SA, and Stones W. Anion gap as a prognostic tool for risk stratification in critically ill patients - a systematic review and meta-analysis [J]. BMC Anesthesiol, 2016, 16(1):68. DOI:10.1186/s12871-016-0241-y.

[6] Lee SB, Kim DH, Kim T, et al. Anion gap and base deficit are predictors of mortality in acute pesticide poisoning [J]. Hum Exp Toxicol, 2019, 38(2):185–192. DOI:10.1177/0960327118788146.

[7] Mitra B, Roman C, Charters KE, et al. Lactate, bicarbonate and anion gap for evaluation of patients presenting with sepsis to the emergency department: A prospective cohort study [J]. Emerg Med Australas, 2020, 32(1):20–24. DOI:10.1111/1742-6723.13324.

[8] Cheng B, Li D, Gong Y, et al. Serum Anion Gap Predicts All-Cause Mortality in Critically Ill Patients with Acute Kidney Injury: Analysis of the MIMIC-III Database [J]. Dis Markers, 2020, 2020: 6501272. DOI:10.1155/2020/6501272.

[9] Banerjee T, Crews DC, Wesson DE, et al. Elevated serum anion gap in adults with moderate chronic kidney disease increases risk for progression to end-stage renal disease [J]. Am J Physiol Renal Physiol, 2019, 316(6):F1244–F1253. DOI:10.1152/ajprenal.00496.2018.

[10] Leskovan JJ, Justiniano CF, Bach JA, et al. Anion gap as a predictor of trauma outcomes in the older trauma population: correlations with injury severity and mortality [J]. Am Surg, 2013, 79(11):1203–1206.

[11] Yang SW, Zhou YJ, Zhao YX, et al. The serum anion gap is associated with disease severity and all-cause mortality in coronary artery disease [J]. J Geriatr Cardiol, 2017, 14(6):392–400. DOI:10.11909/j.issn.1671-5411.2017.06.008.

[12] Sahu A, Cooper HA, and Panza JA. The initial anion gap is a predictor of mortality in acute myocardial infarction [J]. Coron Artery Dis, 2006, 17(5):409–412. DOI:10.1097/00019501-200608000-00002.

[13] Johnson AE, Pollard TJ, Shen L, et al. MIMIC-III, a freely accessible critical care database [J]. Sci Data, 2016, 3: 160035. DOI:10.1038/sdata.2016.35.

[14] Chen Q, Chen Q, Li L, et al. Serum anion gap on admission predicts intensive care unit mortality in patients with aortic aneurysm [J]. Exp Ther Med, 2018, 16(3):1766–1777. DOI:10.3892/etm.2018.6391.

[15] Allard J, Cotin S, Faure F, et al. SOFA--an open source framework for medical simulation [J]. Stud Health Technol Inform, 2007, 125: 13–18.

[16] Le Gall JR, Lemeshow S, and Saulnier F. A new Simplified Acute Physiology Score (SAPS II) based on a European/North American multicenter study [J]. JAMA, 1993, 270(24):2957–2963. DOI:10.1001/jama.270.24.2957.

[17] Feng M, McSparron JI, Kien DT, et al. Transthoracic echocardiography and mortality in sepsis: analysis of the MIMIC-III database [J]. Intensive Care Med, 2018, 44(6):884–892. DOI:10.1007/s00134-018-5208-7.

[18] Jaddoe VW, de Jonge LL, Hofman A, et al. First trimester fetal growth restriction and cardiovascular risk factors in school age children: population based cohort study [J]. BMJ, 2014, 348: g14. DOI:10.1136/bmj.g14.

[19] Ma Q, Luo Y, Guo P, et al. Clinical effects of Xinmailong therapy in patients with chronic heart failure [J]. Int J Med Sci, 2013, 10(5):624–633. DOI:10.7150/ijms.5779.

[20] Stewart S, MacIntyre K, Hole DJ, et al. More ‘malignant’ than cancer? Five-year survival following a first admission for heart failure [J]. Eur J Heart Fail, 2001, 3(3):315–322. DOI:10.1016/s1388-9842(00)00141-0.

[21] Wang Q, Dong L, Jian Z, et al. Effectiveness of a PRECEDE-based education intervention on quality of life in elderly patients with chronic heart failure [J]. BMC Cardiovasc Disord, 2017, 17(1):262. DOI:10.1186/s12872-017-0698-8.

[22] Kraut JA, and Madias NE. Serum anion gap: its uses and limitations in clinical medicine [J]. Clin J Am Soc Nephrol, 2007, 2(1):162–174. DOI:10.2215/CJN.03020906.

[23] Gabow PA, Kaehny WD, Fennessey PV, et al. Diagnostic importance of an increased serum anion gap [J]. N Engl J Med, 1980, 303(15):854–858. DOI:10.1056/NEJM198010093031505.

[24] Ali Sheikh MS, Salma U, Zhang B, et al. Diagnostic, Prognostic, and Therapeutic Value of Circulating miRNAs in Heart Failure Patients Associated with Oxidative Stress [J]. Oxid Med Cell Longev, 2016, 2016: 5893064. DOI:10.1155/2016/5893064.

[25] Fulop M, Horowitz M, Aberman A, et al. Lactic acidosis in pulmonary edema due to left ventricular failure [J]. Ann Intern Med, 1973, 79(2):180–186. DOI:10.7326/0003-4819-79-2-180.

[26] Zymlinski R, Biegus J, Sokolski M, et al. Increased blood lactate is prevalent and identifies poor prognosis in patients with acute heart failure without overt peripheral hypoperfusion [J]. Eur J Heart Fail, 2018, 20(6):1011–1018. DOI:10.1002/ejhf.1156.

[27] Orn S, and van Hall G. Does a normal peripheral lactate value always indicate an aerobic tissue metabolism? [J]. Eur J Heart Fail, 2017, 19(8):1034–1035. DOI:10.1002/ejhf.863.

[28] Kawase T, Toyofuku M, Higashihara T, et al. Validation of lactate level as a predictor of early mortality in acute decompensated heart failure patients who entered intensive care unit [J]. J Cardiol, 2015, 65(2):164–170. DOI:10.1016/j.jjcc.2014.05.006.

[29] Kotake Y. Unmeasured anions and mortality in critically ill patients in 2016 [J]. J Intensive Care, 2016, 4: 45. DOI:10.1186/s40560-016-0171-2.

